# Peripheral blood transcriptomic profiling indicates molecular mechanisms commonly regulated by binge-drinking and placebo-effects

**DOI:** 10.1101/2023.03.21.23287501

**Authors:** Amol Carl Shetty, John Sivinski, Jessica Cornell, Lisa Sadzewicz, Anup Mahurkar, Xing-Qun Wang, Luana Colloca, Weihong Lin, Maureen A. Kane, Chamindi Seneviratne

**Author notes:** Correspondence: Chamindi Seneviratne, MD, 670 W. Baltimore Street, Institute for Genome Sciences, HSF3, Baltimore, Maryland, MD21201.

## Abstract

Molecular changes associated with alcohol consumption arise from complex interactions between pharmacological effects of alcohol, psychological/placebo context surrounding drinking, and other environmental and biological factors. The goal of this study was to tease apart molecular mechanisms regulated by pharmacological effects of alcohol - particularly at binge-drinking, from underlying placebo effects. Transcriptome-wide RNA-seq analyses were performed on peripheral blood samples collected from healthy heavy social drinkers (N=16) enrolled in a 12-day randomized, double-blind, cross-over human laboratory trial testing three alcohol doses: Placebo, moderate (0.05g/kg (men), 0.04g/kg (women)), and binge (1g/kg (men), 0.9g/kg (women)), administered in three 4-day experiments, separated by minimum of 7-day washout periods. Effects of beverage doses on the normalized gene expression counts were analyzed within each experiment compared to its own baseline using paired-t-tests. Differential expression of genes (DEGs) across experimental sequences in which each beverage dose was administered, as well as responsiveness to regular alcohol compared to placebo (i.e., pharmacological effects), were analyzed using generalized linear mixed-effects models. The 10% False discovery rate-adjusted DEGs varied across experimental sequences in response to all three beverage doses. We identified and validated 22 protein coding DEGs potentially responsive to pharmacological effects of binge and medium doses, of which 11 were selectively responsive to binge dose. Binge-dose significantly impacted the Cytokine-cytokine receptor interaction pathway (KEGG: hsa04060) across all experimental-sequences that it was administered in, and during dose-extending placebo. Medium dose and placebo impacted pathways hsa05322, hsa04613, and hsa05034, in the first two and last experimental sequences, respectively. In summary, our findings add novel, and confirm previously reported data supporting dose-dependent effects of alcohol on molecular mechanisms and suggest that the placebo effects may induce molecular responses within the same pathways regulated by alcohol. Innovative study designs are required to validate molecular correlates of placebo effects underlying drinking.

## INTRODUCTION

Alcohol misuse is the third-leading risk factor for premature death and disability in the U.S., and fifth worldwide^1^. Alcohol permeates to virtually all tissues in the body, resulting in significant alterations in organ function through direct or indirect effects leading to multisystemic pathophysiological consequences linked to over 200 health conditions creating a substantial global disease-burden on the society^1,2^. Pathophysiological consequences of alcohol use/misuse differ based on various characteristics of drinking such as the amount, frequency, chronicity, and the type of alcoholic beverage. The pharmacodynamics of these varying characteristics of alcohol consumption are orchestrated by numerous modulating factors such lifestyle or biological factors like the genetic makeup, age, gender, microbiome composition, and etc. ^3-7^.

Recent large-scale genome-wide association studies (GWAS) of alcohol consumption patterns have identified multiple genetic loci and pathways associated with drinking^8-10^. However, the genetic variation often influences traits through altering expression levels of the involved genes via cis- or trans-acting regulation. The genotype-based modulation of gene expression is further fine-tuned by various interacting environmental exposures (i.e., exposome) and epigenomic elements. Thus, gene expression profiling, particularly at a transcriptome-wide analysis (TWA), offer an effective tool to capture a snapshot into the dynamic and context-dependent molecular responses arising from simultaneous or concurrent interplay of various biological and external factors. Because of complexity in specimen collection and prohibitive cost of TWA, gene expression analyses in response to alcohol use/misuse in living humans are often conducted with selective genes based on research hypotheses^11,12^. Apart from the published animal or *in vitro* studies ^13-15^, a recent study by Marvomatis et al. utilized genetic data from a large population-based GWAS to impute transcriptomic associations with alcohol consumption patterns, leveraging cis-acting genetic variants^16^. A large-scale study such as this has the advantage of mitigating some of the variability in gene expression amongst individuals. Nonetheless, because gene expression is a complex and non-linear product resulting from many interacting biological and contextual factors, a comprehensive understanding of effects of drinking behavior on alterations in gene expression requires study paradigms that incorporate individual responses that are experimentally measured while assessing the interacting exposome.

Studies deciphering mechanisms underlying *Drinking* behavior have demonstrated that at least some of the effects of alcohol are accounted for by non-pharmacological component driven by psychological phenomena such as placebo effects. Placebo beverage administration studies conducted in laboratory settings have repeatedly demonstrated that, the individuals who were presented with a placebo beverage often believed the beverages to contain alcohol^17,18^. These observations are further corroborated by slower attentional processing^18^, subjective measures of intoxication^17^ and increased craving for alcohol^19^ seen in response to placebo alcohol administration. However, despite the long history of studies administering placebo alcohol beverages, it is unclear whether the behavioral and subjective outcomes are induced by or stem from molecular-level changes similar to, or distinct from consumption of regular alcohol. Such knowledge may not only improve our understanding of physiological or pathological mechanisms, but also help develop tools to potentially manipulate molecular mechanisms to enhance non-pharmacological responses to improve diagnosis and treatment of alcohol misuse.

In the present study, we performed a TWA using peripheral blood samples collected from a cohort of binge drinkers who were enrolled in a cross-over human laboratory trial conducted in a controlled environment specifically designed to identify biomarkers of binge drinking, while considering placebo effects underlying drinking behavior^20^. Binge drinking is one of the most common patterns of alcohol misuse that increases the risk of developing alcohol use disorder AUD and other deleterious health consequences of alcohol misuse^21^. According to the 2019 National Survey on Drug Use and Health (NSDUH), approximately 24% of the U.S. population aged 12 years and older reported at least one binge drinking episode during the past month, surpassing the population diagnosed with an alcohol use disorder (AUD)^21^. Here, we specifically investigated whether there are, (1) molecular mechanisms commonly regulated by placebo effects and consumption of regular alcohol, (2) pathways or molecular mechanisms that are regulated by binge-level alcohol consumption, but not by lower amounts of drinking, and (3) genes that are potentially regulated by pharmacological effects of alcohol.

## METHODS

### Participants

This study was conducted with a subset of the population analyzed in a parent study that sought to validate serotonin transporter (SERT) mRNA as a quantitative biomarker of binge alcohol consumption in the absence of AUD and pharmacological or behavioral treatments ^20^. Briefly, participants were binge drinking, adult healthy volunteers of Hispanic or non-Hispanic European ancestry. A binge drinking episode was defined as five or more (men) or four or more (women) standard drinks in the past 30 days (one standard drink = 14g of pure alcohol) in one sitting (Robbins et al., 2020)). A detailed list of inclusion and exclusion criteria were reported previously ^20^.

### Study Design

Enrolled participants were randomized to receive placebo and two alcohol doses in counterbalanced order, balancing for sequence of treatment (i.e., beverage dose) and gender within subgroups, in a double-blind human laboratory study. The three doses were: (1) placebo, (2) 0.5g/kg (men) or 0.4g/kg (women) alcohol (medium-dose), and (3) 1g/kg (men) or 0.9g/kg (women) alcohol that correspond to binge drinking conditions (binge-dose). Each beverage dose was given in separate, but otherwise identical four-day long experiments. The beverage free starting day of each experiment was used as the baseline (D0) for each experiment, and the remaining three days involved 2-hour beverage administration sessions where participants received an identical dose each day within an experiment (Exp). Beverage dose differed between the three experiments. Therefore, each participant was scheduled to receive three beverage doses in the three separate experiments, each of which consisted of three once-daily sessions of an identical dose, randomly assigned for that specific experiment. A minimum of seven days living in the community separated experiments allowing for washout periods of more than five half-lives (t_1/2_) of median human cell mRNA (t1/2=10h) ^22^ and alcohol (t1/2= 4-5h) ^23^. Participants were closely monitored and not allowed to eat or drink anything that was not part of the standardized protocol. A total of 24 mL of whole blood was collected daily from each participant using collection tubes containing acid citrate dextrose (ACD) buffer (Vacutainer®, Becton-Dickinson, Franklin Lakes, NJ) at baseline and 17.5 to 18h after the end of each drinking session to allow for late-onset gene expression alterations ^24-26^.

### Measures of known direct biomarkers of alcohol consumption

We measured two known biomarkers, breath alcohol concentration (BrAC) and ethyl glucuronide (EtG) at each session day within an experiment to objectively assess the presence of alcohol and its metabolite EtG at baseline, pre- and post-drinking. The BrAC measurements were collected at admission, immediately prior to and after each dosing session, as well as immediately prior to blood sample collection for RNA and EtG analyses. The WBC and plasma derived from the same whole blood samples were used for total RNA extractions and the detection of EtG, respectively. The EtG levels were determined using liquid chromatography tandem mass spectrometric method as described previously ^20,27,28^.

### Total RNA isolation and sequencing

For this retrospective transcriptomic analysis, we used white blood cell (WBC) samples isolated from whole blood collected at D0 and the final day of each experiment (i.e., after three daily drinking sessions (D3). Total RNA was extracted from WBC using Macherey-Nagel’s NucleoSpin® miRNA kit and RNA/DNA Buffer kit (Takara Bio USA, Inc. Doral, Fl, USA) according to manufacturer’s guidelines. The RNA integrity was tested using an Agilent bioanalyzer system, and all samples that had an RNA integrity number (RIN) greater than seven were used for sequencing. Paired end libraries were prepared and sequenced using Illumina HiSeq4000 platform (Illumina, Inc., San Diego, CA) at a sequencing depth of 150 million reads at 100 bp PE length sequences.

### Sequencing data analyses

The raw sequence reads generated for each sample were analyzed using the CAVERN analysis pipeline ^29^. The FastQC toolkit was used to assess read quality for downstream analyses. The reads were aligned with the human reference genome GRCh38 (Ensembl repository) using fast splice-aware aligner HISAT2 ^30^ under default parameters to generate the alignment BAM files. The read alignments were assessed to compute gene expression counts for each gene using the HTSeq count tool ^31^ and the human reference annotation (GRCh38). The raw read counts were normalized for library size and dispersion of gene expression and utilized in following downstream analyses at individual gene and pathway levels.

### Influence of beverage doses on individual genes

As we detected significant sequence effects on SERT mRNA expression levels in the parent study (i.e., the SERT mRNA expression levels differed significantly when the same beverage dose was administered in Exp-1 vs. Exp-2 vs. Exp-3)^20^, we assessed differential expression of genes (DEGs) between D0 and D3 within each dose-by-experiment group separately using DESeq2, rather than averaging expression levels across the three experiments for a given dose. We analyzed DEGS for the following nine conditions comparing expression levels between D0 and D3 paired data from each individual: (1) placebo administered in Exp-1; (2) medium-dose in Exp-1; (3) binge-dose in Exp-1; (4) placebo in Exp-2; (5) medium-dose in Exp-2; (6) binge-dose in Exp-2; (4) placebo in Exp-3; (5) medium-dose in Exp-3; (6) binge-dose in Exp-3. The p-values were generated using the Wald test implemented in DESeq2 and then corrected for multiple hypothesis testing using the Benjamini–Hochberg correction method ^32^. Next, we explored the possibility of assessing the effects of each dose on baseline-adjusted fold-changes across all three experiments where a certain dose was administered, by combining experiment-specific data via a generalized linear mixed-effects model that used individual as random and experiment and beverage dose as fixed effects adjusting for the sequences in which the beverage doses were administered. The expression levels of genes are a cumulative response to many environmental, biological, and psychological factors as stated above. Hence, to tease apart pharmacologic effects of alcohol from potential placebo effects on DEGs and pathways, we performed the following contrasts within each experiment: (1) binge-dose vs. placebo, and (2) medium-dose vs. placebo. Next, we combined the gene expression levels from the contrasts across the three experimental sequences using a generalized linear mixed-effects model similar to the exploratory analysis performed within each dose group. The significant DEGs between conditions in all comparisons were determined using a false discovery rate (FDR) of 10% and a minimum absolute log2 fold-change of 0.6.

### Pathway analyses

We analyzed the impact of administration of the three beverage doses on molecular pathways using iPathwayGuide (Advaita Bioinformatics, Plymouth Michigan, USA) that identifies impact of DEGs within pathways defined by the Kyoto Encyclopedia of Genes and Genomes (KEGG; Release 100.0+/11-12, Nov 21) ^33,34^ based on, (1) over representation of DEGs within a pathway, and (2) perturbation of the pathway computed by the measured expression changes propagating along the pathway topology. Perturbations are computed using gene ontologies obtained from the Gene Ontology Consortium database (2021-Nov4) ^35^, network of regulatory relations from BioGRID, and the Biological General Repository for Interaction Datasets v4.4.203. Oct. 25th, 2021 ^36^. Two independent probability values, pORA (over-representation P-value) and pAcc (total accumulation P-value), are combined to calculate a unique pathway-specific P-value using Fisher’s method to assess overall pathway “impact”. Putative mechanisms were inferred using *Advaita Knowledge Base* (AKB v1910, www.advaitabio.com) when measured gene expression changes were consistent with the computed sequence of events within a pathway. We performed pathway analyses on all contrasts analyzed for effects at single gene level listed above (i.e., 20 analyses in total (nine analyses for dose effects within each experiment: six analyses for the dose-vs-placebo effects within each experiment; three analyses for each dose across its’ three experiments; two analyses for the dose-vs-placebo effects across its’ three experiments)). The iPathwayGuide input files consisted of all genes with measured expression levels, and pathways were identified with DEGs unadjusted for FDR at p<0.05 statistical threshold, and the pathway-specific P-values were subsequently adjusted for FDR of 10%.

### Gene Expression validation with NanoString assays

Protein-coding genes that were found to be differentially expressed between regular alcohol (binge or medium doses) and placebo with RNA-seq were validated using customized *NanoString nCounter* assays (NanoString Technologies, Seattle, WA, USA). The validation sample included 27 pre- and post-treatment total RNA sample pairs (i.e., 54 samples in total) with a 71% overlap with the discovery cohort and were profiled concurrently. The NanoString code set additionally included two housekeeping genes (GAPDH, and MPP1) for data normalization, and spike-in positive (N=6) and negative (N=8) controls to set a minimum threshold count above background for data analyses. Capture and reporter probe sets were hybridized with total RNA samples (100 ng minimum per sample) and the hybridization reactions were loaded on an *nCounter Prep Station* (Version 1, NanoString Technologies, Seattle, WA, USA) for removing excess capture and reporter probes, immobilize and align hybridized complexes for binding to cartridges to create an image for single molecular counting and data collection, as per the manufacturer’s guidelines.

#### NanoString Data analysis

The mRNA raw data counts were analyzed using nSolver™ Analysis Software (Version 4.0; NanoString Technologies, Seattle, WA, USA). Six negative controls were used to perform background thresholding. Positive controls were used to perform technical normalization to adjust any lane-by-lane variability due to differences in hybridization or binding. After technical normalization, the NanoString readings were analyzed using the same mixed model analyses to compare expression levels between binge-dose vs placebo and medium-dose vs. placebo beverage administrations.

## RESULTS

We used WBC samples from 16 heavy social drinkers who were enrolled in a 12-day human laboratory trial, where they received three different doses of alcohol beverages that they consumed within 2h sessions as described in methods section. Whole transcriptomic data was generated with RNA-seq from peripheral blood WBC collected at D0 and D3 from each participant in three experiments. The numbers of participants assigned to dose-by-experiment categories are presented in Table 1 along with participant characteristics. Transcriptomic analysis resulted in 28,294 Ensembl annotated genes (43.97% of all Ensembl annotated genes). All participants had zero BrAC readings at admission to each 4-day long in-house experiments, at the time of blood draws on D0 and D3, and immediately prior to the start of drinking sessions (Fig.1; consort diagram is presented in Cornell et al.^20^). The average BrAC readings taken immediately following the 2-h drinking sessions differed significantly between dose groups, both in the discovery (p = 6.86E-08) and the validation (p = 2.68E-06) cohorts (Table 1). At the time of blood draws (17-18h post-drinking sessions) for transcriptomic analyses, the EtG levels were above lower limit of quantification (LLOQ) in majority of participants after consuming the binge-dose, and undetectable (below LLOQ) after the placebo dose.

**Fig. 1:**
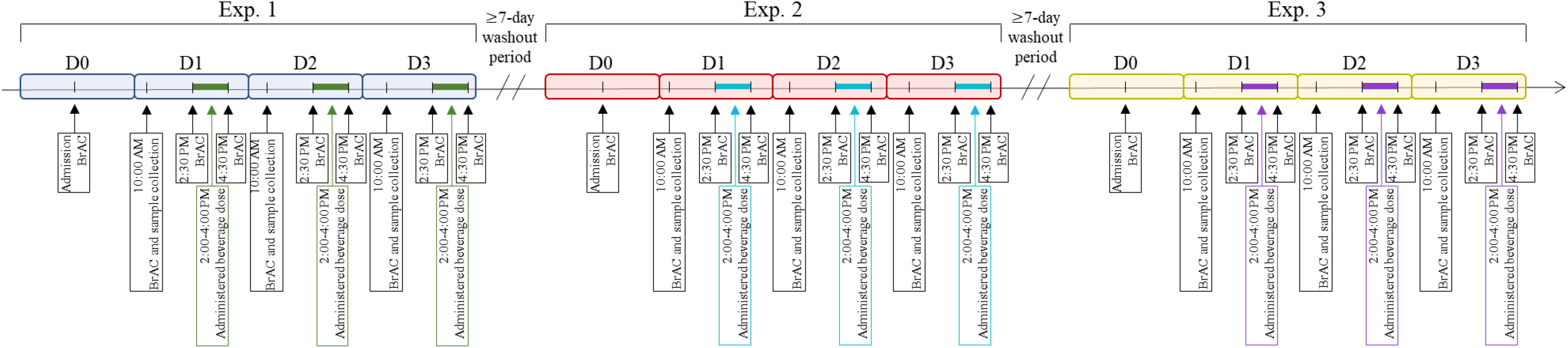
Scheduled dose administration, BrAC readings, and sample collection for all experiments. The three doses were assigned to Exp.1-3 randomly as detailed previously^20^. Exp.1-3 = sequentially conducted experiments 1 through 3. D0-3 = session days within each experiment. BrAC= breath alcohol concentration. Colored horizontal bars within D1-3 represent 2h-drinking sessions. The arrow across Exp1-3 indicate the direction of experimental sequence.

**Table 1:** Demographics and study characteristics. **Footnote:** ns= P >0.05 for the comparison of the three groups and comparison between placebo and medium or high dose alcohol groups. P-values were derived from Fisher Exact test for categorical variables and Kruskal Wallis test for continuous variables. *Calculated at baseline of each experiment. **Calculated using standard drinks consumed in the 30 days prior to initial in-person screen. #Number of participants who used other drugs (excluding alcohol) in the 90 days prior to initial in-person screen. ##Breath alcohol content (BrAC) reading units were g/210 L.

| Characteristic | Total Population | Placebo | Medium | High | P_value |
| --- | --- | --- | --- | --- | --- |
| **Discovery cohort** |  |  |  |  |  |
| Total No. of sample pairs | 31 | 13 | 11 | 7 | ns |
| Exp.1 |  | 5 | 3 | 2 |  |
| Exp.2 |  | 3 | 4 | 4 |  |
| Exp.3 |  | 5 | 4 | 1 |  |
| Age | 27.50 | 27.38 | 29.09 | 30.70 | ns |
| Gender (% male) | 68.75 | 61.54 | 81.82 | 85.70 | ns |
| Average BMI (SD)* | 24.71 (3.69) | 24.57 (3.62) | 25.28 (3.56) | 25.46 (3.58) | ns |
| Average AUDIT score (SD) | 8.56 (2.48) | 8.69 (2.66) | 8.64 (2.16) | 9.25 (2.12) | ns |
| Past 30 days drinking measures** |  |  |  |  |  |
| Average drinks per drinking day (SD) | 4.6 (1.9) | 4.6 (2.0) | 4.5 (1.2) | 4.2 (1.5) | ns |
| Average number binge episodes (SD) | 7.3 (4.9) | 7.7 (5.2) | 7.9 (4.4) | 8.6 (5.9) | ns |
| Past 90 days other drug use<sup>#</sup> |  |  |  |  |  |
| Nicotine (no.) | 2 | 2 | 0 | 1 |  |
| Marijuana (no.) | 6 | 5 | 2 | 2 |  |
| Cocaine (no.) | 0 | 0 | 0 | 0 |  |
| Average BrAC Levels<sup>##</sup> |  |  |  |  |  |
| Pre-Dose, Day 3 (SD) | 0.000 (0) | 0.000 (0) | 0.000 (0) | 0.000 (0) | ns |
| Post-Dose, Day 3 (SD) | 0.045 (0.051) | 0.001 (0.002) | 0.047 (0.021) | 0.106 (0.051) | 6.86E-08 |
| EtG levels |  |  |  |  |  |
| ≥LLOQ (%), Baseline | 0 (0) | 0 (0) | 0 (0) | 0 (0) |  |
| 0>LLOQ (%), Baseline | 2 (6.45) | 1 (7.69) | 0 (0) | 1 (14.29) |  |
| ≥LLOQ (%), Post-Dose | 7 (22.58) | 0 (0) | 1 (9.09) | 6 (85.71) |  |
| 0>LLOQ (%), Post-Dose | 3 (9.68) | 0 (0) | 2 (18.18) | 1 (14.29) |  |
| **NanoString Validation cohort** |  |  |  |  |  |
| Total No. of sample pairs | 28 | 10 | 11 | 7 |  |
| Exp.1 |  | 5 | 4 | 2 |  |
| Exp.2 |  | 2 | 4 | 4 |  |
| Exp.3 |  | 3 | 3 | 1 |  |
| Age | 26.38 | 24.80 | 26.72 | 27.29 | ns |
| Gender (% male) | 68.75 | 60.00 | 81.82 | 85.71 |  |
| Average BMI (SD)* | 25.64 (3.12) | 25.16 (3.02) | 25.88 (3.31) | 24.79 (3.38) | ns |
| Average AUDIT score (SD) | 8.69 (2.09) | 8.50 (2.07) | 8.00 (1.48) | 8.57 (2.44) | ns |
| Past 30 days drinking measures** |  |  |  |  |  |
| Average drinks per drinking day (SD) | 4.9 (1.7) | 4.8 (1.9) | 4.9 (1.4) | 4.6 (1.9) | ns |
| Average number binge episodes (SD) | 7.4 (4.3) | 7.7 (5.0) | 7.5 (4.5) | 7.9 (6.3) | ns |
| Past 90 days other drug use<sup>#</sup> |  |  |  |  |  |
| Nicotine (no.) | 4 | 3 | 2 | 2 |  |
| Marijuana (no.) | 7 | 5 | 5 | 4 |  |
| Cocaine (no.) | 1 | 0 | 1 | 0 |  |
| Average BrAC Levels<sup>##</sup> |  |  |  |  |  |
| Pre-Dose, Day 3 (SD) | 0.000 (0) | 0.000 (0) | 0.000 (0) | 0.000 (0) | ns |
| Post-Dose, Day 3 (SD) | 0.046<br>(0.047) | 0.000<br>(0) | 0.059<br>(0.034) | 0.094<br>(0.038) | 7.43E-06 |
| EtG levels |  |  |  |  |  |
| ≥LLOQ (%), Baseline | 1 (3.57) | 0 (0) | 1 (9.09) | 0 (0) |  |
| 0>LLOQ (%), Baseline | 2 (7.14) | 1 (10) | 0 (0) | 1 (14.29) |  |
| ≥LLOQ (%), Post-Dose | 8 (28.57) | 0 (0) | 3 (27.27) | 5 (71.43) |  |
| 0>LLOQ (%), Post-Dose | 1 (3.57) | 0 (0) | 0 (0) | 1 (14.29) |  |

### Alcohol dose-associated responses in WBC gene expression

Differential expression (i.e., fold changes between D3 and D0) for all measured genes in response to the nine dose-by-experimental-sequence categories are presented in Table S1. We adjusted the P-values across all analyses for multiple testing, using false discovery rate (FDR) with a q-value threshold of 0.1 indicating significance, as suggested by Van den Oord and Sullivan ^37^. The numbers of DEGs differed both across experiments (i.e., the same dose administered in different experimental sequences) and across doses (Fig. 2). We did not detect any DEGs that were common to all three experiments in response to any of the three doses at the transcriptome-wide threshold, further implying sequence effects of dose administration. However, as shown in Fig. 2, five significant DEGs were detected in at least two of the three experiments in which the binge-dose was administered (Fig. 2F). These were: In Exp-1 and 3, Ring Finger Protein 182 (RNF182; upregulated in both) and MHC Class-I Polypeptide-Related Sequence-A (MICA; downregulated in Exp-1 and upregulated in Exp-3); Fos Proto-Oncogene, AP-1 Transcription Factor Subunit (FOS), Nicotinamide Phosphoribosyltransferase (NAMPT), and Dual Specificity Phosphatase 1 (DUSP1) upregulated in both Exp-1 and 2. Charcot-Leyden Crystal Galectin (CLC) gene was significantly upregulated in both binge dose and placebo treated experiment 1 (Fig. 2A). TMSB4X Pseudogene 1 (TMSB4XP1) was significantly upregulated in binge dose treated Exp-1 but downregulated in placebo treated Exp-1 with a less than 1.5-fold change. In addition to these DEGs, there were 12, 49, and 221 significant DEGs in at least one of the three placebo-, medium- and binge-dose treated experiments, respectively (Fig. 2A-F; Table S1). When the baseline-adjusted gene expression levels in response to binge and medium doses were compared with the gene expression levels in response to placebo within the same experimental sequence (i.e., Fig. 2G-I), a fewer number of significant DEGs were detected in both dose categories across experimental sequences. A complete list of fold changes and P-values for the dose vs. placebo comparisons within each experiment are presented in Table S1.

**Fig. 2:**
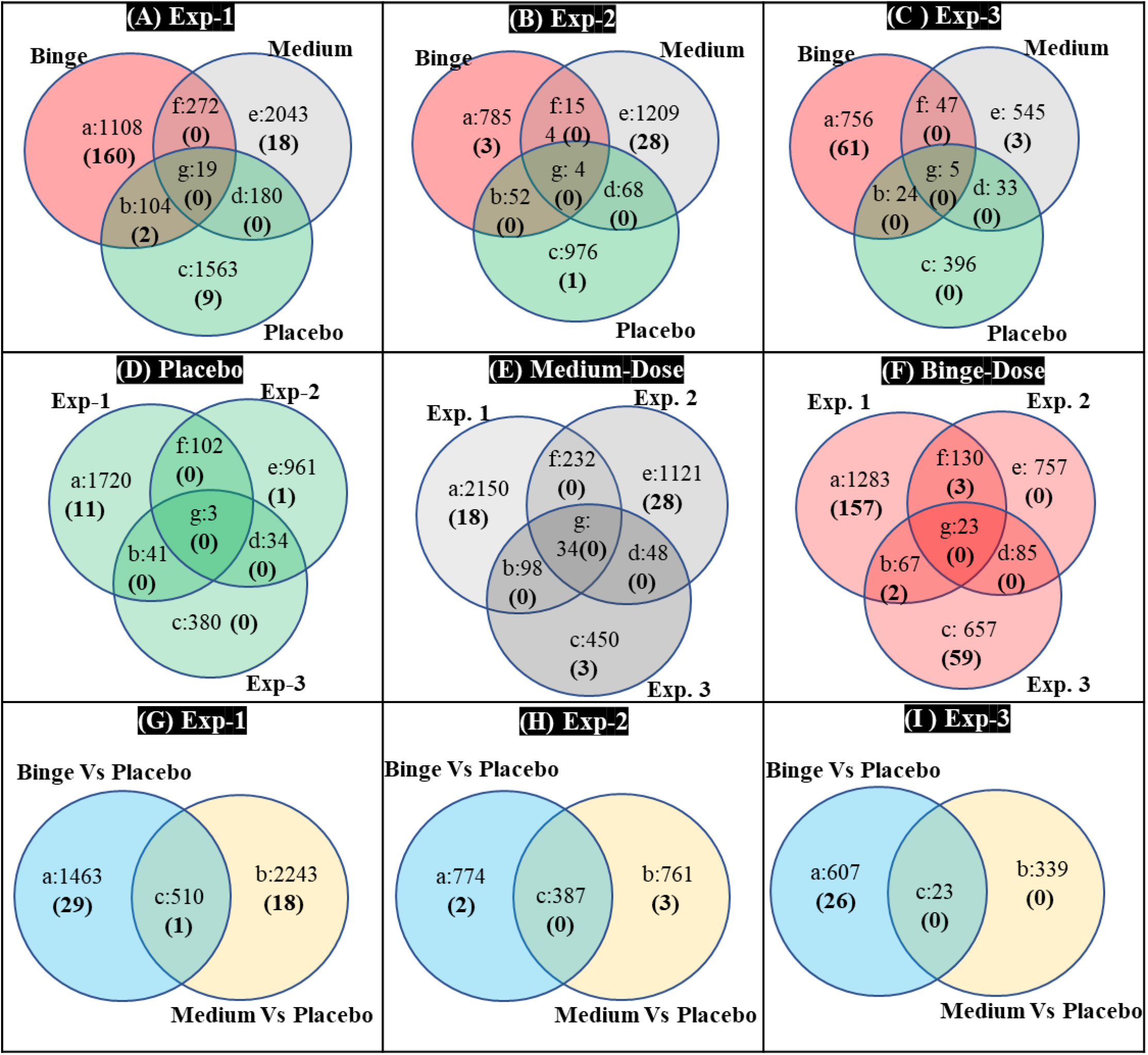
DEGs across experiments in response to beverage doses detected with RNA-seq. Numbers outside parentheses represent the numbers of significant DEGs (p<0.05) prior to FDR adjustments. Numbers within parentheses represent numbers of DEGs that remained significant after adjustments for FDR. **Fig. 2A-C** illustrate numbers of DEGs detected within each of the three beverage dose groups stratified by the three experimental sequences that they were administered in, across all participants. **Fig. 2D-F** illustrate the distribution of detected DEGs within each experimental sequence when stratified by the beverage doses assigned to an experiment, across all participants. **Fig. 2G-I** illustrate the distribution of detected DEGs detected in dose-vs-placebo comparisons performed within each experimental sequence when stratified by the beverage doses assigned to an experiment, across all participants.

When the data across experimental sequences for each dose were combined using a mixed model, there were 54 significant DEGs in response to binge dose including the above-mentioned genes RNF182, FOS, NAMPT, and DUSP1. There were 17 significant DEGs in response to medium dose and only one significant DEG in response to the placebo (Guanosine Monophosphate Reductase 2; GMPR2) in the combined analysis (Table S1).

### Alcohol dose-associated responses in WBC gene expression excluding potential placebo responses

The mixed model analyses of RNA-seq data across all three experiments comparing binge-dose with placebo (binge-vs-placebo) or medium-dose with placebo (medium-vs-placebo) revealed a total of 36 and 64 significant DEGs (FDR adjusted), respectively. The distribution of DEGs in the binge-vs-placebo comparison were, 72.73% protein coding, 18.18% processed pseudogenes, 6.06% antisense RNA genes, and 3.03% sense-intronic genes. The distribution of DEGs in the medium-vs-placebo comparison were, 53.85% protein coding, 11.54% pseudogenes, 19.23% anti-sense RNA genes, 5.77% sense-intronic, 3.85% lincRNA, 1.92% miRNA and 3.85% other genes. Table 2 lists all significant protein coding DEGs that were assessed with NanoString nCounter assays in both dose-vs-placebo comparisons. As shown in Table 2, *NanoString* assays validated the RNA-seq findings for 19 out of 52 protein coding DEGs in binge-vs-placebo comparison group (36.53%), and 11 out of 52 DEGs in medium-vs-placebo group (21.15%). Twenty-three DEGs that were selectively detected in the medium-vs-placebo group with RNA-seq, were found to be significant in binge-vs-placebo group with the NanoString assays. Four and 22 significant DEGs detected with RNA-seq in binge-vs-placebo and medium-vs-placebo groups, respectively, failed validation with *NanoString*. Overall, compared to RNA-seq, *NanoString* assays detected more significant DEGs in binge-vs-placebo group (24 vs. 43 out of 52 tested genes) and fewer significant DEGs in medium-vs-placebo group (28 vs. 17 out of 52 tested genes).

**Table 2:**
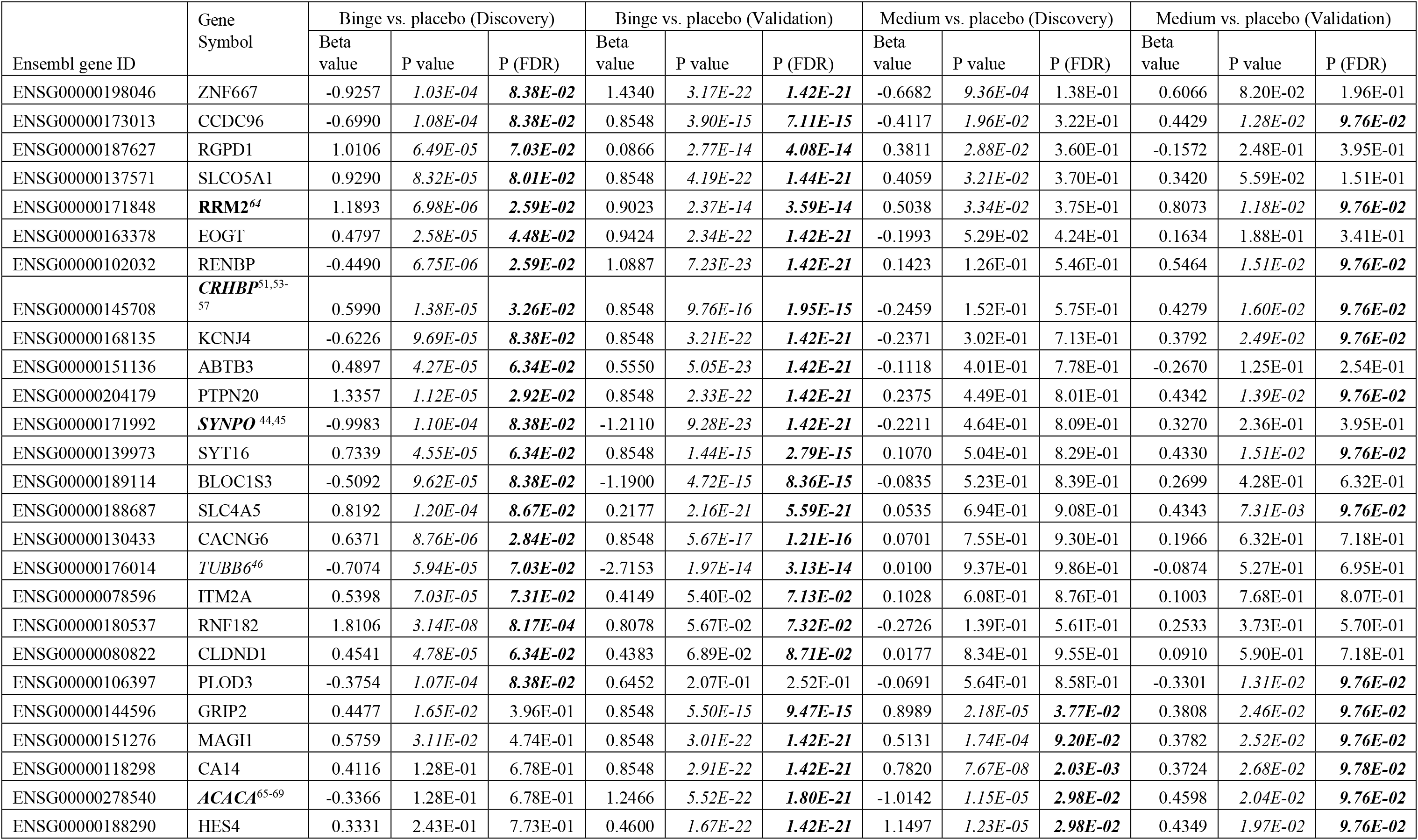
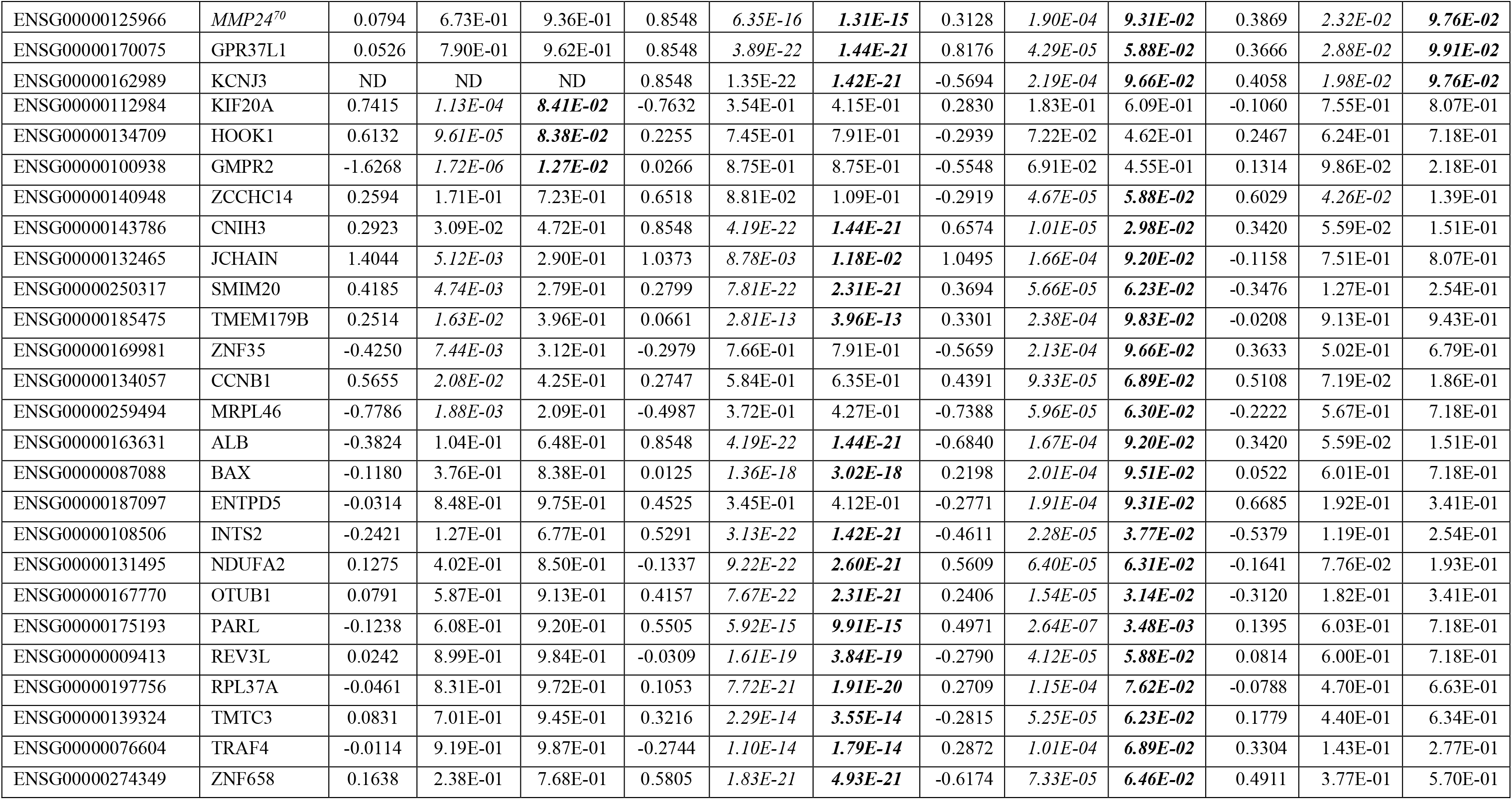
Significantly differentially expressed protein-coding genes identified and validated in dose-vs-placebo comparisons across experiments. **Footnote**: ND = not detected. Italicized values represent significant P-values prior to FDR adjustments and bold italicized values represent P-values that remained significant after adjustments for FDR. Bolded and italicized gene symbols - previously reported in human and animal studies; bolded gene symbols - previously reported only in human studies; italicized gene symbols - previously reported only in animal studies.

### The most impacted pathways overlapped across beverage doses

A total of 341 pathways were identified in the nine dose-by-experiment categories (Table S2). Of these, ten pathways were significantly (FDR adjusted) impacted by at least one of the three tested doses based on the number of enriched genes and their perturbations within a pathway (Table S3). Table 3 summarizes four significantly impacted pathways that were detected in more than one of the three experiments for any dose tested in this study. As presented in Table 3, the FDR adjusted P-values were significant for all four pathways in response to placebo when administered in Exp-3 (i.e., when the placebo was administered in the last double-blind experiment after completing two experiments in which the participants received regular alcohol at the binge and medium doses). The KEGG pathways *Systemic lupus erythematosus* (hsa05322), *Neutrophil extracellular trap formation* (hsa04613), and *Alcoholism* (hsa05034) were impacted by placebo via upregulation of seven out of all 80 genes in H2A, H2B, H3 and H4 gene classes (H2BC21, H2BC5, H2AC16, H2BC4, H2AC8, H4C15, and H3C13) expressed in the nucleosome (GO:0000786; P-value (FDR) = 0.002). Upregulated genes H2BC21, H2BC5, and H2AC16 interacted to form a network through which the placebo putatively impacted hsa05322, hsa04613, and hsa05034. Within the putative network, H2BC21 and H2BC5 were predicted to interact via activation/catalyzation, while upregulated genes H2BC5 and H2AC16 were predicted to interact with each other through binding of their protein products. These interactive predictions were further supported by the finding that the seven genes, along with three additional genes (CEACAM6, IKBKG, and TAF4B; total of 10) were annotated to protein heterodimerization activity (GO:0046982; 10 out of all 259 genes - P-value (FDR) = 0.023).

**Table 3:** Significantly altered pathways detected within each 4-day experiment in response to beverage doses. **Footnote:** Italicized values represent significant P-values prior to FDR adjustments and bold italicized values represent P-values that remained significant after adjustments for FDR.

| Pathway Name<br>(KEGG ID) | Dose | Exp-1 |  |  |  | Exp-2 |  |  |  | Exp-3 |  |  |  |
| --- | --- | --- | --- | --- | --- | --- | --- | --- | --- | --- | --- | --- | --- |
|  |  | Count<br>(DE/All) | Rank | Unadjusted<br>P-value | Adjusted<br>P-value | Count<br>(DE/All) | Rank | Unadjusted<br>P-value | Adjusted<br>P-value | Count<br>(DE/All) | Rank | Unadjusted<br>P-value | Adjusted<br>P-value |
| Systemic lupus<br>erythematosus<br>(hsa05322) | Binge | 9/94 | 37 | 0.3171 | 0.8260 | 4/94 | 34 | 0.1914 | 0.8042 | 23/94 | 3 | 4.60E-06 | **0.0004** |
|  | Medium | 23/97 | 3 | 0.0003 | **0.0372** | 17/98 | 1 | 2.7646E-05 | **0.0073** | 8/98 | 1 | 0.0038 | 0.6920 |
|  | Placebo | 10/100 | 5 | 0.0152 | 0.8386 | 5/101 | 13 | 0.0524 | 0.8718 | 7/101 | 1 | 0.0002 | **0.0251** |
| Neutrophil<br>extracellular trap<br>formation<br>(hsa04613) | Binge | 17/157 | 11 | 0.0200 | 0.5607 | 5/157 | 49 | 0.7031 | 0.8980 | 24/157 | 1 | 8.56E-07 | **0.0001** |
|  | Medium | 35/160 | 1 | 8.39E-06 | **0.0018** | 19/161 | 2 | 0.0003 | **0.0456** | 9/161 | 2 | 0.0353 | 0.6920 |
|  | Placebo | 11/164 | 11 | 0.0688 | 0.9808 | 3/165 | 19 | 0.4896 | 0.8718 | 8/165 | 2 | 0.0004 | **0.0318** |
| Cytokine-cytokine receptor<br>interaction<br>(hsa04060) | Binge | 27/199 | 1 | 0.0004 | **0.0987** | 18/199 | 2 | 0.0008 | **0.0953** | 7/199 | 38 | 0.3566 | 0.8035 |
|  | Medium | 26/207 | 29 | 0.2997 | 0.8852 | 19/207 | 7 | 0.0299 | 0.7199 | 7/207 | 7 | 0.3464 | 0.7626 |
|  | Placebo | 16/203 | 7 | 0.0250 | 0.9808 | 11/203 | 14 | 0.1171 | 0.8718 | 4/203 | 4 | 0.0014 | **0.0538** |
| Alcoholism<br>(hsa05034) | Binge | 15/141 | 23 | 0.0990 | 0.8133 | 3/140 | 41 | 0.3866 | 0.8228 | 22/140 | 2 | 9.40E-07 | **0.0001** |
|  | Medium | 27/143 | 4 | 0.0006 | **0.0494** | 18/143 | 3 | 0.0027 | 0.2340 | 8/143 | 3 | 0.0211 | 0.6920 |
|  | Placebo | 12/148 | 9 | 0.0342 | 0.9808 | 3/148 | 23 | 0.9537 | 0.9826 | 7/148 | 3 | 0.0009 | **0.0448** |

The H2A, H2B, H3 and H4 gene classes were also upregulated in response to the binge dose in Exp-3 (21 genes) and medium dose in Exp-1 (20 genes). However, unlike by placebo where the pathways hsa05322, hsa04613, and hsa05034 were altered exclusively via the upregulation of H2A, H2B, H3 and H4 gene classes, additional gene classes contributed to upregulation of hsa05322, hsa04613, and hsa05034 KEGG pathways when binge or medium doses were administered. All genes that contributed to altered pathways are listed in Table S3.

The Cytokine-cytokine receptor interaction pathway (KEGG: hsa04060) was the most significantly impacted pathway in response to the binge-dose in Exp-1, Exp-2 (see Table 3), and combined analysis of all binge-dose administered experiments (FDR adjusted P-value = 0.004; Fig. 3A). As shown in Fig. 3A and 2B, the impact of binge dose on pathway hsa04060 was due to the presence of many DEGs (21 out of 201 genes) contributing to the pathway, as well as the propagation of signals (red lines) from extracellular chemokines (CXC subfamily), and IL6/12-like and IL17-like cytokines. The sequences of pathway signals for measured expression levels in response to binge-dose were consistent with the computed sequence of events for following genes inferring two putative networks within hsa04060 (Fig. 3C). Conversely, in response to placebo, hsa04060 was impacted during Exp-3 via four DEGs (IL2, IL11, MSTN, and CXCR6; Table S2), but the measured expression levels were not consistent with the computed sequence of events (i.e., gene-by-gene interactions resulting in putative mechanisms were not identified within the pathway), implying a relatively weaker impact of the placebo on has04060.

**Fig. 3:**
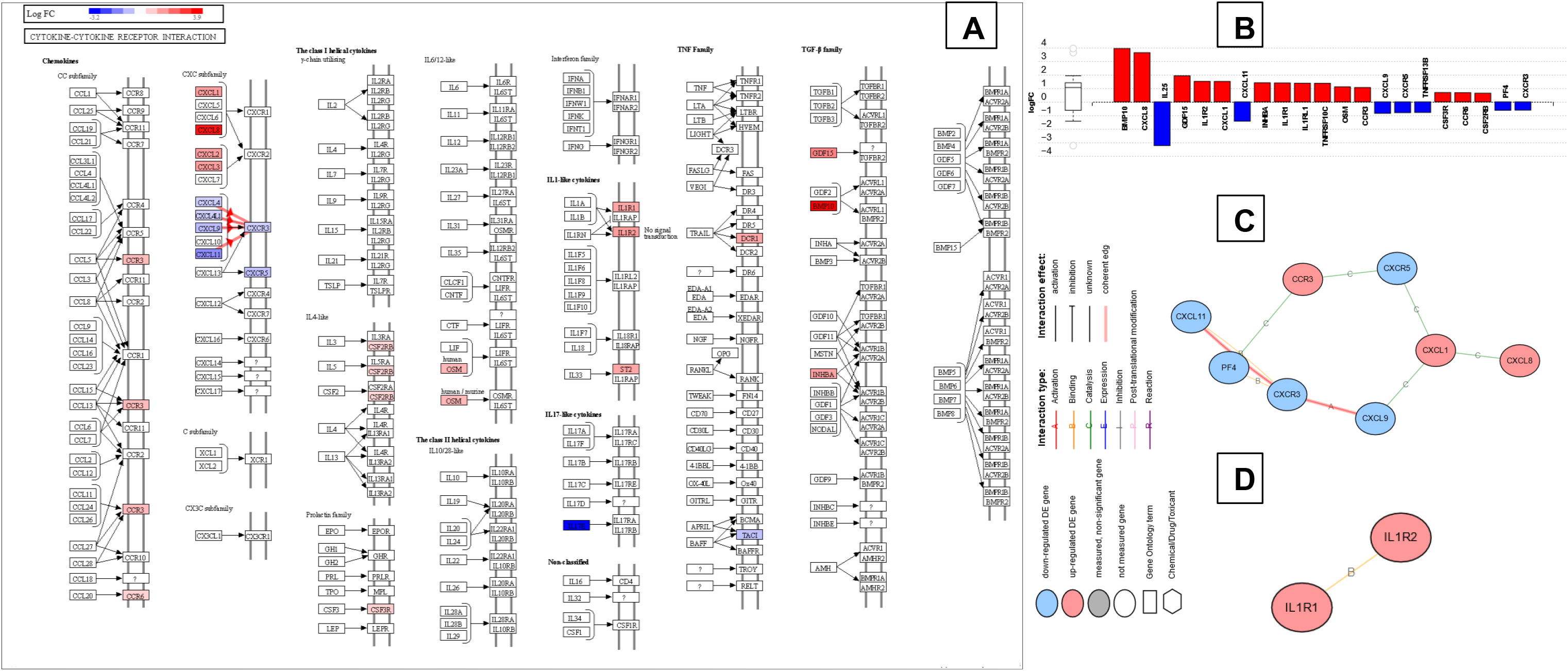
Binge drinking and placebo alcohol were associated with alterations in the Cytokine-cytokine receptor interaction pathway (hsa04060) Upregulated genes are shown in red, and downregulated genes are shown in blue in all figures. **(A)** The pathway diagram overlayed with the computed perturbation of each gene within pathway hsa04060 in the mixed model analysis of all binge-dose administered visits. The perturbations account both for the genes’ measured fold changes and the accumulated perturbations propagated from any upstream genes (accumulation). The highest negative perturbation is shown in dark blue, while the highest positive perturbation in dark red. The legend describes the values on the gradient. For legibility, one gene may be represented in multiple locations in the diagram and one box may represent multiple genes in the same gene family. A gene is highlighted in all locations it occurs in the diagram. For each gene family, the color corresponding to the gene with the highest absolute perturbation is displayed. Red lines with arrows indicate sequence of steps and the direction of the signal propagation (i.e., coherent cascades) for which the observed expression changes are agree with the expected changes. **(B)** Measured expression levels of genes within pathway hsa04060 that correspond to **Fig. 3A**, ranked based on their absolute values of log fold change. The box and whisker plot on the left summarizes the distribution of all the differentially expressed genes in pathway hsa04060. The box represents the 1^st^ quartile, the median and the 3^rd^ quartile, while the outliers are represented by circles. **P**utative mechanisms (Fig 2C and 2D) through which binge-dose may act on the genes measured to be differentially expressed in pathway hsa04060. **Fig. 3C** represent the types of gene-by-gene interactions within the cascade illustrated with red arrows in **Fig. 3A. Fig 3D** represents upregulated IL-1 and IL-2 interactions within IL-1–type cytokine sub pathway, suggesting a proinflammatory response.

The *iPathwayguide* analyses did not identify any significantly impacted pathways when placebo responses were subtracted from binge- and medium-dose responses (i.e., in binge-vs-placebo and medium-vs-placebo comparisons).

## DISCUSSION

The present study represents an initial attempt to identify molecular-level responses to binge-drinking and underlying placebo. In a transcriptome-wide analysis (TWA) of peripheral blood samples collected from 16 healthy binge drinkers of European ancestry who were enrolled in a crossover, double-blind oral alcohol administration paradigm, we demonstrate commonly and distinctly regulated genes and pathways by alcohol and placebo. Furthermore, the sequence of dose administration had a variable effect on gene expression alterations and the pathways they formed. Specifically, while placebo induced more DEGs in the first experimental sequence than in the latter, they had significant impact on molecular pathways when administered in the last experiment (i.e., following binge and medium doses). Conversely, binge and medium doses showed significant effects at single-gene and pathway level starting from the first experimental sequence. Together, these findings indicate dose-extending placebo effects at a molecular level.

At the single gene level, two DEGs - RNF182 (ring finger protein 182) and CACNG6 (calcium voltage-gated channel auxiliary subunit gamma 6), were found to be upregulated by both RNA-seq and NanoString assays, selectively in the binge-dose administered experiments regardless of adjustments for placebo responses. This observation suggests strong pharmacological effects on their regulation at binge-level drinking. None of the other 250 binge-dose responsive (FDR adjusted) DEGs remained significant when the placebo responses were subtracted. The RNF182 encodes a ubiquitin ligase that enables ubiquitin-protein transferase activity in the cytoplasm resulting in inhibition of innate immune responses triggered by TLR^38^, and expressed commonly in the brain, and bone marrow. The CACNG6 encodes an integral membrane protein that forms one of the five subunits (gamma subunit) of voltage-gated L-type calcium ion channels that stabilizes the calcium channel in its inactive or closed state^39^. Direct effects of alcohol on RNF182 or CACNG6 expression levels are not reported in literature. However, Peng et al., reported a SNP (rs2500086) associated with alcohol-induced affective symptoms during alcohol withdrawal in a cohort of European Americans, that was in eQTL with RNF182 ^40^. Similarly, previous studies have demonstrated that alcohol transiently inhibit L-type calcium ion channels at intoxicating concentrations^41,42^. Thus, it is possible that binge drinking may inhibit L-type calcium ion channels through upregulation of CACNG6, as we have detected in this study.

Ten other protein-coding DEGs were detected with RNA-seq and validated with NanoString assays, that were selectively responsive to the binge-dose (but not the medium-dose) only when their expression levels were compared with that of the placebo group (ZNF667, RGPD1, SLCO5A1, EOGT, ABTB3, SYNPO, BLOC1S3, TUBB6, ITM2A, and CLDND1). Of these, the greatest expression fold changes were seen with SYNPO, TUBB6 and SLCO5A1. Protein product of SYNPO (synaptopodin) is involved in formation of cytoskeleton of dendritic spines and podocytes^43^. The TUBB6 (Tubulin B6) is also involved in the formation of synaptic cytoskeleton regulating neurotransmission. Both SYNPO and TUBB6 were reported previously to be associated with response to alcohol, including upregulation in nucleus accumbens of P rats^44,45^ and mice^46^. Our findings indicated a downregulation of SYNPO and TUBB6, and the reversal in direction could perhaps be related to the differences in molecular adaptations to alcohol in our sample of heavy social drinkers compared to those with an AUD modeled in the rodent studies. The SLCO5A1 belongs to the superfamily of organic anion transporting polypeptides (also known as Oatps/OATPs) which are amphipathic transporters that mediate cellular drug and xenobiotic influx^47,48^, including renal secretion of acamprosate^49^. The SLCO5A1 mRNA are abundantly expressed in human peripheral blood cells including PBMCs, monocytes, immature dendritic cells^50^, but the effects of alcohol on SLCO5A1 expression levels or its protein product (OAT5) are not known. Our data suggest an upregulation of peripheral blood cell SLCO5A1 in response to the binge-dose compared to placebo.

Another eight protein-coding DEGs were detected in response to binge- and medium dose groups each (or possibly both doses as all 16 were detected to be significant with NanoString assays), when compared to the placebo group (Table 2). The fact that these associations were not significant when unadjusted for placebo responses suggest that their expression may be modulated by placebo-associated contextual factors that an individual experiences while drinking. For example, the widely studied and well-replicated corticotropin releasing hormone-binding protein gene (CRHBP) that mediates neuroendocrine stress response, was found to be upregulated in our study in response to binge-dose (by both RNA-seq and NanoString) and possibly the medium dose (detected only with *NanoString*) when adjusted for placebo responses, but not in response to either of the alcohol doses or the placebo alone. These findings agree with the study by Haass-Koffler et al that demonstrated a downregulation of CRHBP gene in the center nucleus of the amygdala associated with reduced ethanol consumption in ethanol preferring rats^51^, and human studies that showed CRHBP to be significantly associated with stress-induced phenotypes of alcohol^52^, including AUD comorbid with anxiety^53^, alcohol use/misuse^54^, negative affect and the negative consequences of drinking^55^, comorbid AUD in schizophrenic patients^56^, and greater levels of subjective tension and negative mood^57^. Although we have not systematically assessed stress levels in our cohort during alcohol administration procedure, placebo effects have been shown to moderate physiological and subjective stress responses through associated contextual factors^58,59^.

There are several notable observations in our pathway-level analyses: (1) significantly altered pathways were identified when responses to beverages were analyzed separately, but not when placebo responses were discounted from binge- or medium dose responses; (2) placebo alcohol impacted the pathways significantly associated with binge- and medium doses when administered in the last (i.e., 3rd) of the sequentially scheduled experiments a participant underwent, and (3) at the least in our study, placebo responsive genes within these commonly impacted pathways were fewer compared to the numbers of genes altered by regular alcohol consumption. These findings suggest a potentially significant placebo component (representing the context of beliefs) altering molecular mechanisms underlying drinking. Our cross-over study design included washout periods that lasted a minimum of 7 days between dose-specific experiments. Then we assessed dose-dependent gene expression alterations by normalizing post-drinking expression data within an experiment by its’ own baseline. Furthermore, objective measures of plasma EtG levels and BrAC readings at baselines of each experiment confirmed that the participants were abstinent from alcohol at admissions prior to alcohol administration procedure. Hence, it is possible that the significant responses detected when placebo was administered in the last experiment, were partly due to conditioning by repeated administrations of regular alcohol in prior experiments. In fact, placebos have been shown to successfully extended analgesic effects of opioids in the management of pain^60,61^. Here, we specifically detected four pathways suggestive of alcohol dose-extended effects of placebo: Three pathways via differential expression of seven histone genes (H3C13, H2AC16, H4C15, H2AC8, H2BC4, H2BC5, and H2BC21; Table 3) and the cytokine-cytokine receptor interaction pathway via IL2, IL11, MSTN, and CXCR6. Of note, the cytokine-cytokine receptor interaction pathway was also highly significant in response to binge-dose. The studies exploring the role of placebo conditioning on molecular pathways have demonstrated that plasma interleukin (IL)-2 was reduced in placebo conditioned immunosuppression^62,63^. Whether the downregulation of IL-2 detected in our study leads to immune suppression remains to be explored in a more comprehensive approach.

There are a few caveats that should be considered when interpreting our findings. First, we had a modest sample size that was vulnerable to imbalances in genetic effects regulating confounding gene expression (not assessed in the present analysis) between dose-by-experiment categories. However, unlike in a parallel group design, this study was strengthened by its cross-over design that randomly assigned participants to three sequences, each of which were 4-days long inpatient experiments that allowed each participant to have their own baseline gene expression measurements prior to the administration of a beverage dose. Consequently, we were able to use baseline adjusted gene expression counts in all between group analyses improving the confidence in our results. Furthermore, as previously reported^20^, the study was conducted in a highly controlled setting maintaining environmental factors constant across participants and experiments as much as possible. Second, it would have been ideal to validate all the nominally significantly associated DEGs detected with RNA-sequencing, prior to inclusion in pathway analyses. Alternatively, because of the prohibitively expensive cost, we limited our validation analyses with NanoString to assessing TWA-significant protein-coding DEGs detected in the comparisons between regular alcohol doses and placebo that potentially captured pharmacologic effects of alcohol. Therefore, the non-coding DEGs detected in dose-vs-placebo comparisons, as well as the pathway associations in response to dose-by-experiment categories should be interpreted with caution. Third, the subjective effects of placebo alcohol administration were not assessed systematically, limiting our ability to directly correlate behavioral constructs with the molecular alterations. Further, we used the commercially available non-alcoholic beer (O’Douls) as the blinded placebo which had a similar consistency and aroma to the regular alcohol beverage administered in the study. While this approach is in line with published behavioral studies that explored placebo effects underlying drinking^17^, it is still possible that the non-alcoholic contents may have contributed to the detected gene expression alterations either directly or indirectly acting upon other physiological systems. Even if this was the case, the DEGs that we detected to be associated with pharmacological effects of regular alcohol would likely survive as the main difference between the two beverage types was the content of alcohol which was negligible in the non-alcoholic beer, as evidenced by the zero breathalyzer readings immediately after drinking and below detection-level EtG values in the placebo arm at the time of blood draw for expression analyses (Table 1). Whether the DEGs associated with the non-alcoholic beverage were in fact due to true placebo effects need further exploration using a beverage-free arm akin to no-treatment arms in placebo studies^60^, and by incorporating newer technologies such as virtual reality that simulate drinking environments. Despite these shortcomings, our study presents the first transcriptome-wide assessment of placebo alcohol administration, providing a framework for more structured studies in the future.

In conclusion, we present initial clues of molecular mechanisms commonly regulated by pharmacological effects of alcohol and placebo effects underlying drinking. Important next steps would be to explore whether these molecular mechanisms can be optimized with improved study paradigms and applying more sensitive molecular techniques such as single-cell transcriptomics or profiling plasma cell-free transcriptome to uncover non-invasive (i.e., peripheral) biomarkers for the identification of novel treatment targets and diagnostics.

## Data Availability

All data produced in the present study are available upon reasonable request to the authors following publication of the peer-reviewed article

## DATA AVAILABILITY

Full transcriptomic data used to support the findings of this study will be deposited in NIH Gene Expression Omnibus (GEO) repository (https://www.ncbi.nlm.nih.gov/geo/) and will be embargoed until results from full datasets are in press.

## ACKNOWLEDGEMENTS

The research was supported by the NIAAA grants K23AA020899 and R01AA026291 (to CS), “micro-grants (vouchers)” to CS through the UMB Clinical Translational Research Initiative (ICTR program) that partially facilitated human laboratory procedures through CTSA grant 1UL1TR003098, and in part by the University of Maryland Baltimore, School of Pharmacy Mass Spectrometry Center (SOP1841-IQB2014) to MAK. Authors thank Sandra Ott and Holy Bowen at the Maryland Genomics center affiliated with the UMSOM/IGS for their assistance with the *NanoString* assays.

